# Predicting chronic kidney disease progression using small pathology datasets and explainable machine learning models

**DOI:** 10.1101/2024.04.08.24305414

**Authors:** Sandeep Reddy, Supriya Roy, Kay Weng Choy, Sourav Sharma, Karen M Dwyer, Chaitanya Manapragada, Bahareh Nakisa

## Abstract

Chronic kidney disease (CKD) poses a major global public health burden, with approximately 7 million affected. Early identification of those in whom disease is likely to progress enables timely therapeutic interventions to delay advancement to kidney failure. This study developed explainable machine learning models leveraging pathology data to accurately predict CKD trajectory, targeting improved prognostic capability even in early stages using limited datasets. Key variables used in this study include age, gender, most recent estimated glomerular filtration rate (eGFR), mean eGFR, and eGFR slope over time prior to incidence of kidney failure. Supervised classification modelling techniques included decision tree and random forest algorithms selected for interpretability. Internal validation on an Australian tertiary centre cohort (n=706; 353 with kidney failure and 353 without) achieved exceptional predictive accuracy, with the area under the receiver operating characteristic curve (ROC-AUC) reaching 0.94 and 0.98 on the binary task of predicting kidney failure for decision tree and random forest, respectively. To address the inherent class imbalance, centroid-cluster-based under-sampling was applied to the Australian dataset. To externally validate the performance of the model, we applied the model to a dataset (n=597 adults) sourced from a Japanese CKD registry. To overcome risks of overfitting on small sample sizes, transfer learning was subsequently employed by fine-tuned machine learning models on 15% of the external dataset (n=89) before evaluating the remaining 508 patients. This external validation demonstrated performant results with an ROC-AUC of 0.88 for the decision tree and 0.93 for the random forest model. Decision tree model analysis revealed the most recent eGFR and eGFR slope as the most informative variables for prediction in the Japanese cohort, aligning with the underlying pathophysiology. The research highlights the utility of deploying explainable machine learning techniques to forecast CKD trajectory even in the early stages utilising limited real-world datasets.

## Introduction

Approximately 7 million people were living with chronic kidney disease (CKD) globally in 2017 (1). CKD is defined as a reduction in the kidney filtration rate and/or the presence of proteinuria for 3 months or more. CKD is traditionally classified according to stages, with Stage 1/2 being early-stage, Stage 3/4 mid-stage and Stage 5 kidney failure. In Australia, there are an estimated 1.7 million aged 18 and over with CKD (2, 3). Whilst most people with CKD are living in early (63.9%) and mid (34.6%) stage kidney disease, there has been a more than doubling of people requiring kidney replacement therapy (in the form of dialysis or transplantation) between 2000 – 2020 to over 27,000 Australians, which is predicted to increase a further 30% by 2030(4). The early identification of people who are likely to progress to kidney failure presents an opportunity to deliver targeted interventions to alter the trajectory of the disease. Indeed, early detection and management can prevent progressive CKD by as much as 50%, a figure that may be even higher in the era of new renoprotective therapies (5, 6). The healthcare, societal and economic implications are profound if kidney failure can be delayed or prevented. The annual cost of CKD was estimated to be $9.9 billion in Australia in 2021, with upwards of $1.2 billion absorbed by dialysis alone (7). For the individual, time without kidney failure is associated with a higher quality of life, and maintenance of day-to-day activities including the ability to work.

CKD is diagnosed with a combination of a blood test to assess biochemical kidney function and a urine test to determine the amount of protein in the urine (3, 8). The data arising is used to determine the Stage of CKD categorizing the condition from stage 1 (mild damage) to stage 5 (kidney failure), which is essential for determining the appropriate management and treatment strategies. Despite the simplicity of these investigations, proteinuria data is often missing, compromising optimal care delivery (9). Furthermore, the vast number of factors that may influence disease progression are not captured with these two tests alone. Currently, predicting who is likely to progress to kidney failure is based on clinical acumen underpinned by the trajectory of kidney disease. There is a clinical need to be able to identify those individuals who are at greatest risk of progressive disease and kidney failure by integrating other relevant data sets.

The Kidney Failure Risk Equation (KFRE) is a risk prediction equation that estimates the risk of progression to kidney failure for an individual with chronic kidney disease (CKD) at 2 and 5 years (10). It has been internationally validated, including in Australia, and is in clinical use in some jurisdictions (11). Essential variables in the equation include age, sex, eGFR, and urine albumin/creatinine ratio. Serum albumin, bicarbonate, phosphorus, and adjusted calcium are optional parameters. However, the equation has important limitations in that it requires a urine albumin/creatinine ratio, which is infrequently done in clinical practice, applies only to later stages of CKD (G3-G5) and considers only kidney failure (requiring dialysis) as the outcome (9). Furthermore, the KFRE is a static equation which does not take multiple readings and variation of variables over a time into account.

Recent advancements in artificial intelligence (AI) and machine learning have shown promise in improving the prediction of CKD progression compared to traditional risk equations like the KFRE (12-14). AI models can analyse complex, longitudinal data and uncover non-linear relationships between variables, potentially leading to more accurate and personalised risk predictions (8, 15). Ferguson and colleagues developed a random forest model to predict the progression of chronic kidney disease (CKD) using patient demographics and over 80 different laboratory variables, such as complete blood cell counts, chemistry panels, and urine analysis (16). This retrospective study aimed to predict a 40% decline in eGFR or kidney failure using data from a single time point. The model achieved a high area under the curve (AUC) in both internal testing (0.88 at 2 years and 0.84 at 5 years) and external validation (0.87 at 2 years and 0.84 at 5 years). Whilst the interrogation of routine laboratory data is advantageous, the sheer number of variables limits its clinical application. Aoki and colleagues developed a machine learning model applicable across all stages of CKD (12).

Their retrospective observational study conducted using laboratory data involved 110,264 adults over 5 years. The model utilised random forest survival methods with a 7-variable risk classifier. It accurately predicted a significant decline in eGFR (>30%) within 5 years, with an AUC-ROC of 0.85. Remarkably, it performed consistently across diverse demographic groups, with an AUC range of 0.83-0.88, and relied on standard pathology results, enhancing its applicability and cost-efficiency. The model highlighted the eGFR slope as the most significant predictor of progression, a novel insight compared to prior studies. Other key variables included initial eGFR, urine albumin-to-creatinine ratio, serum albumin, age, and sex. The study’s unique approach using a machine-learning random forest survival model allowed for a nuanced analysis of time-based variables, offering an advantage over traditional regression methods by accommodating complex, non-linear data. Numerous other studies have demonstrated the efficacy of longitudinal analysis, underscoring the limitations of KFRE as a static model (8, 13, 15). By training models with time-series data, there is potential to uncover complex, non-linear relationships between variables without relying on opaque methods like deep neural networks, where interpretability is often a significant challenge.

Previous studies on CKD prediction and prognosis using ML techniques have emphasized the impact of dataset characteristics and variable selection on model performance. However, in these studies, there is mostly a lack of external validation on independent datasets, which is crucial for assessing generalizability. The use of neural networks as a baseline model is also unexplored. Many previous models for CKD prognosis have a “black box” nature, hindering clinical adoption due to poor interpretability. XAI techniques like SHapley Additive explanations (SHAP) and Local Interpretable Model-agnostic Explanations (LIME) improve model transparency and trustworthiness, bridging the gap between ML predictions and clinical applicability. In addition to XAI techniques, counterfactual analysis emerges as a pivotal method, slightly modifying input data points to observe changes in the model’s predictions and providing tangible and actionable insights into the model’s decision-making process.

In order to develop an explainable longitudinal machine learning model that could effectively identify those people with chronic kidney disease (CKD) who would ultimately progress to kidney failure, we carried out a retrospective analysis utilising existing pathology data that was routinely stored in the electronic medical record systems at the Department of Pathology at Northern Health in Melbourne, Australia over the 5-year period spanning from January 1st, 2019 to December 31st, 2023. Upon receiving ethics approval after thorough procedures to ensure patient privacy and anonymity were in place, we were provided secured access to approximately 10,000 eligible patient records that met the key inclusion criteria of being adult patients aged 18 years and over at the time of diagnosis with a baseline estimated glomerular filtration rate (eGFR) of ≥15 mL/min/1.73m^2^ and ≤ 59 mL/min/1.73m^2^ . Kidney failure was defined when two eGFR readings of <15 ml/min/1,73m^2^ were recorded.

Biochemistry arising from dialysis centres were excluded. The electronically extracted records contained basic non-identifiable patient details such as age and gender that were routinely stored for standard clinical pathology purposes and allowed the research team to assign a unique de-identified study number to each patient to further uphold confidentiality principles. Additionally, the Northern Health co-investigator on our study team ensured that all relevant data points were properly anonymised by removing any potential personal identifiers before recording select information into a secure spreadsheet. The secure spreadsheet was developed a priori specifically for the purposes of this analysis based on the identification of key variables and data fields required to answer our research question on predictive modelling from a thorough review of similar previously published studies in leading nephrology journals. The study obtained ethics approval from the Austin Health Human Research Ethics Committee: HREC/100996/Austin-2023.

Our study introduces several novelties in predicting kidney disease using longitudinal data analysis and diverse algorithms, including machine learning algorithms like Decision Trees and Random Forest, and deep learning algorithms like Recurrent Neural Networks (RNNs). The research aimed to identify key variables that predict the progression to kidney failure in patients with CKD, using a three-variable based model that does not require further information such as proteinuria. In a unique methodological advancement, the study employed transfer learning for external dataset validation, enhancing the generalizability and robustness of the predictive models. The study also introduced a technique for handling small datasets by utilizing centroid clustering-based under-sampling to balance the data while maintaining the integrity of the original dataset’s representation, significantly improving the accuracy of kidney failure predictions. Further to this, the novelty of this study lies in its focus on explainable AI (XAI) techniques, such as SHAP, LIME, and counterfactual analysis, to address the “black box” limitation of previous CKD prognosis models. By improving model transparency and interpretability, these methods bridge the gap between ML predictions and clinical applicability, enabling healthcare professionals to understand the rationale behind the model’s decisions. Counterfactual analysis provides actionable insights by identifying the smallest changes needed to alter a prediction, thus informing targeted interventions and personalized treatment plans for enhanced patient care.

## Methodology

In this section, the proposed approach to forecast the CKD trajectory using both the Australian and an external validation dataset is presented. There are four main tasks: Data Preparation, Feature Engineering and Under-sampling, Model Training and Transfer Learning.

### Data Preparation

The Australian dataset was collected internally, with 149,491 patients being assessed as eligible. To validate the model performance, a Japanese dataset consisting of 1,138 newly visiting stage G2–G5 CKD patients was utilised (17). The dataset comprised demographic, biochemical, and clinical data of patients identified with CKD (eGFR values between 15-59mL/min/1.73m^2^). The distinctive characteristics included gender and age, eGFR calculated using the 2009 CKD Epidemiology Collaboration (CKD-EPI) creatinine equation without the race-based coefficient, Serum/LDL/HDL cholesterol, triglycerides, albumin-adjusted calcium, potassium, sodium, phosphate, ferritin, creatinine, urate, C-reactive protein (CRP), 25-hydroxyvitamin D, haemoglobin and haemoglobin A1c (HbA1c) and Urinary albumin-to-creatinine ratio, urinary protein-to-creatinine ratio. Follow-up duration data was also extracted. A minimum of three eGFR measurements per subject was required. For the Australian cohort, patients who had two eGFR readings on the same day after their first eGFR reading falling below 15 mL/min/1.73m^2^ were considered to be on dialysis and excluded. The final number of patients was 10,064 and 597 for the Australian and Japanese cohorts, respectively. The threshold for defining kidney failure was set for patients who exhibited at least two eGFR readings below 15 mL/min/1.73m^2^ with an interval of at least 30 days. In the context of the Japanese dataset, these readings were inherently spaced six months apart. Consequently, the final cohorts comprised 10,064 patients (353 with kidney failure) in the Australian cohort and 597 patients (162 with kidney failure) in the Japanese cohort.

For the Japanese Dataset, the age at each eGFR measurement was adjusted to the nearest whole number based on the exact timing of the test reading since only the age at the initial test was provided. Notably, the eGFR values in the Japanese cohort were initially calculated using the Modification of Diet in Renal Disease (MDRD) study equation, while the Australian data utilised the Chronic Kidney Disease Epidemiology Collaboration (CKD-EPI) equation for eGFR calculation. The eGFR was therefore re-computed for the Japanese Dataset to ensure consistency with the Australian Dataset. Starting from the MDRD-based eGFR, the serum creatinine (SCr) concentrations were reverse-calculated, factoring in the updated age and gender of the patients. The derived SCr values were then converted from the units of mg/dL to µmol/L. Using these standardised SCr values, along with the patient’s age and gender, the eGFR was recalculated according to the CKD-EPI formula. Additionally, the categorical variable for gender was encoded. Figure 1 outlines the patient selection protocol from Australian and Japanese cohorts for CKD prediction.

**Figure 1.**
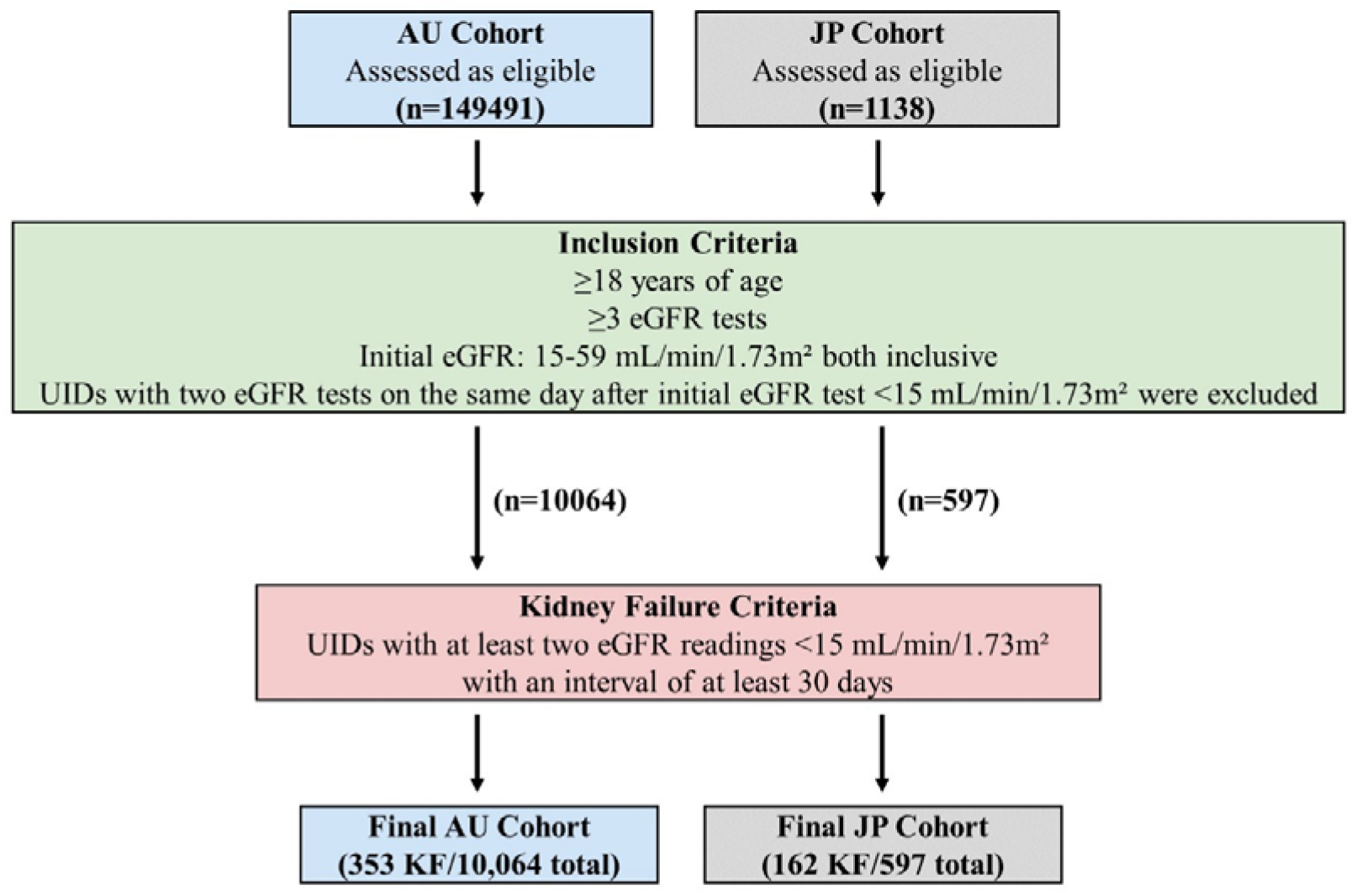
Data preparation for CKD prediction for Australian and Japanese patient cohorts.

A comparison of the Australian and Japanese cohorts is provided in Table 1.

**Table 1.** Summary Statistics for Australian (before under-sampling) and Japanese CKD Datasets.

| Variables | AU Dataset | JP Dataset |
| --- | --- | --- |
| Total number of patients | 10,064 | 597 |
| Total number of records | 154,516 | 3,388 |
| Total number of patients with KF | 353 | 162 |
| Total number of records with KF | 5,543 | 456 |
| Sex (M/F) | 5064/5000 | 435/162 |
| Age (years) | 77.54 ± 12.02 | 71.55 ± 11.47 |
| eGFR (ml/min/1.73m<sup>2</sup>) | 46.64 ± 18.36 | 36.62 ± 14.75 |
| Kidney Failure (%) | 3.58% (5543 out of 154516) | 13.46% (456 out of 3388 records) |

|  | records) |  |
| --- | --- | --- |
| eGFR_slope | $0.17 \pm 1.87$ | $-0.0047 \pm 0.0134$ |
| eGFR_mean (ml/min/1.73m<sup>2</sup>) | $46.64 \pm 15.71$ | $36.62 \pm 13.56$ |
| eGFR_std | $8.78 \pm 4.38$ | $5.22 \pm 3.62$ |

### Feature Engineering

The patient time-series records were truncated prior to the onset of kidney failure, marked by eGFR failing below 15 mL/min/1.73m^2^ for prediction. For tree-based models, a linear regression slope of eGFR values over time was obtained for each patient to capture longitudinal trends in kidney function. Subsequently, the selected features included age, gender, the most recent eGFR value, eGFR time progression slope, and mean eGFR prior to kidney failure onset. The features were normalised with standard scaling for both the Australian and Japanese cohorts.

### Under-sampling (Imbalanced dataset)

The Australian dataset underwent a pre-processing step where centroid-cluster-based under-sampling was applied to address the class imbalance issue, explicitly aiming to balance the majority class (non-kidney failure cases) with the minority class (kidney failure cases). In datasets where one class significantly outweighs the other, models may exhibit a bias towards predicting the majority class, which can result in poor generalisation for instances of the minority class. This method involves clustering the majority class instances with the K-means algorithm and then using the centroids of these clusters as representative samples, effectively decreasing the number of majority class instances. The majority class samples are divided into N number of clusters, where N is the number of minority class samples in the dataset.

This approach helps not only balance the classes but also retain the majority class’s underlying distribution. This methodology has been utilised in small-scale medical datasets, such as a study by Zhang et al. [7], which utilised cluster-based under-sampling to address the diagnosis of breast cancer. Similarly, in a study by Jian et al. [8], this methodology was utilised to address the inherent imbalance challenges in predicting eight different diabetes complications.

Conversely, for the neural network models, specifically Recurrent Neural Networks (RNNs), the slope or mean of the eGFR values were not utilised. Instead, the data was grouped by the patient after normalising the numerical age and eGFR values with min-max scaling and then zero-padded to a fixed sequence length of 50, maintaining consistency before feeding into the RNN. To balance the dataset, TimeSeriesKMeans clustering with Dynamic Time Warping (DTW) was applied to the sequences of the majority class, dividing them into clusters based on the similarity of their time series patterns [8]. Instead of selecting representative sequences from each cluster, the centroids of these clusters—synthetic sequences that represent the “average” pattern within each cluster—are used. The number of clusters was chosen to match the number of instances in the minority class, ensuring that the resulting set of centroid sequences from the majority class equals the size of the minority class. These centroid sequences are then combined with the original sequences from the minority class to create a balanced dataset. The sequential nature within each centroid sequence is preserved due to the time series clustering while addressing class imbalance. The sequential model includes a Simple RNN layer with three units, designed to process input sequences of length 50 representing three features: eGFR values over time, age, and gender. Following this, two Dense layers are utilised: the first layer consists of 10 units with ReLU activation, while the final layer contains a single unit with a sigmoid activation function, enabling the categorisation of the output for binary classification. The model is compiled with the Adam optimiser, set at a learning rate of 0.001 and uses binary cross-entropy as the loss function.

### Model Training

For this study, a model was initially trained and evaluated on the Australian dataset to perform a binary classification task with the objective of predicting kidney failure based on the given features (most recent eGFR, eGFR slope, eGFR mean prior to kidney failure incidence, and age and gender). We employed explainable machine learning algorithms— Decision Tree, Random Forest—and one deep learning algorithm, namely, the Recurrent Neural Networks (RNN), for comparison. Grid search was performed to identify the optimal hyperparameter combination for each algorithm. The model’s evaluation on the Australian dataset was carried out using stratified K-fold (k=5) cross-validation to ensure class balance in each fold [6]. This involved splitting the dataset into five equally sized class-stratified folds. Four of these folds were used for model training, while the remaining fold was used for testing. This process was rotated through all five folds to thoroughly assess the model’s performance. The performance of all classifiers was evaluated using accuracy, precision, recall, specificity, F1 score, and Receiver Operating Characteristic - Area Under the Curve (ROC-AUC) in accordance with the guidelines recommended for reporting results of clinical prediction models (18). The model was fit with the best hyperparameters and retrained on the entire under-sampled Australian dataset.

### Transfer Learning

One of the problems when dealing with small datasets, especially within healthcare datasets, is overfitting. Generally, overfitting occurs when a machine learning model learns not only the underlying patterns in the training data but also its noise and random fluctuations. As a result, while the model may perform exceptionally well on the training data, its performance significantly degrades on new, unseen data because it has essentially memorised the training data rather than learning to generalise from it. Considering the smallness of the datasets utilised in this study, we explored the validation of the performance of our models on an external dataset.

For this purpose, the pre-trained model on the under-sampled Australian dataset set is then evaluated on the Japanese cohort, denoted as the held-out test set. This was done by fine-tuning the model on a small subset (15%) of the Japanese dataset with the same features before being evaluated on the remaining data. Transfer learning is a machine learning method where a model trained on a source task is repurposed as the starting point for a target task [9]. This approach negates the need to train the model from scratch in the target domain, reducing both the need for large training datasets and the time required for training [10]. By utilising a model pre-trained on the source task, transfer learning incorporates patterns from the initial dataset and addresses the common problem of insufficient training data, thereby improving a model’s performance on the target dataset. This approach mitigates the potential for overfitting on the source task and improves generalisation by performing well on an unseen target dataset.

In a basic transfer learning scenario, we define a source domain *D*_*S*_ with data *X*_*S*_ and a task *T*_*S*_, and a target domain *D*_*T*_ with data *X*_*T*_ and a task *T*_*T*_. The goal of transfer learning is to improve the learning of the target predictive function *f*_*T*_*(·)* in *D*_*T*_ using the knowledge from *D*_*S*_ and *T*_*S*_. Typically, a model is initially trained in *D*_*S*_, resulting in parameters θ_*S*_. When adapting this model to *D*_*T*_, we often initialise the target model’s parameters, θ_*T*_, with θ_*S*_(i.e., θ _*T*_^*(0)*^ *=*θ_*S*_), implying that the target model begins with parameters learned from the source model. During fine-tuning, θ_*T*_ is adjusted to better fit the target domain data *X*_*T*_. The updated rule in an iterative training process (like gradient descent) can be represented as:

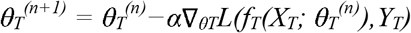

where:

- θ_*T*_^*(n)*^ are the parameters at iteration n,
- α is the learning rate,
- *L* is the loss function that quantifies the discrepancy between the predicted outcomes,
- *f*_*T*_*(X*_*T*_; θ_*T*_^*(n)*^) and the actual values *Y*_*T*_ in the target domain,
- ∇_θ*T*_*L* represents the gradient of the loss function with respect to the parameters θ_*T*_, guiding the update direction.

Figure 2. presents the comprehensive methodology, including the data selection and preprocessing, feature engineering, machine learning (ML) model development on the Australian (AU) cohort, and subsequent transfer learning on the Japanese (JP) cohort for robust cross-validation.

**Figure 2.**
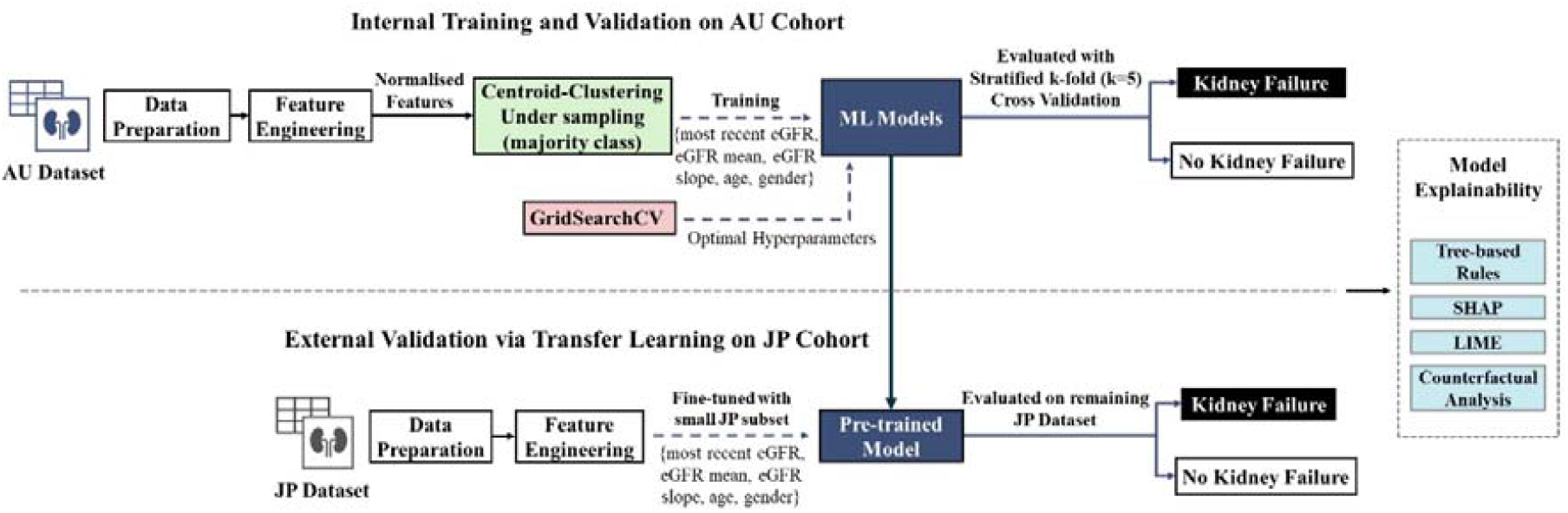
The proposed CKD prediction framework evaluated on both AU and JP datasets, with internal and external validation phases.

## Results

The hyperparameter settings for each classifier are displayed in Table 2. For the decision tree, the model’s hyperparameters are fine-tuned using GridSearchCV and optimised for the ROC-AUC score. The optimal configuration chosen did not incorporate class weight adjustments, and splits were determined by Gini impurity using the ‘best’ method, which evaluated all possible splits at a node to identify the most informative one. The tree nodes are expanded until all leaves are pure or contain fewer than min_samples_split samples, set at 10, to prevent overfitting by ensuring that each decision to divide the data is supported by a sufficient number of samples. Additionally, ‘min_samples_leaf’ is set to 4 and may improve the model’s generalisation by requiring that there be at least a small group of data points that lead to a decision rather than decisions based on single points.

**Table 2.**
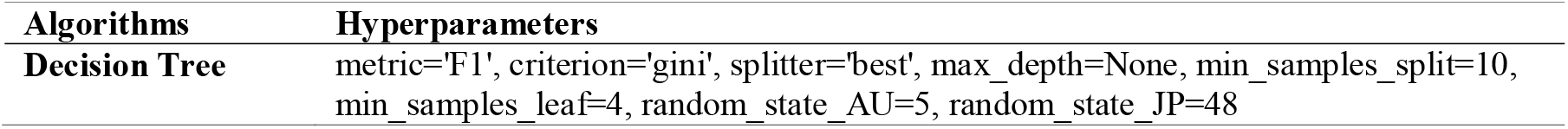
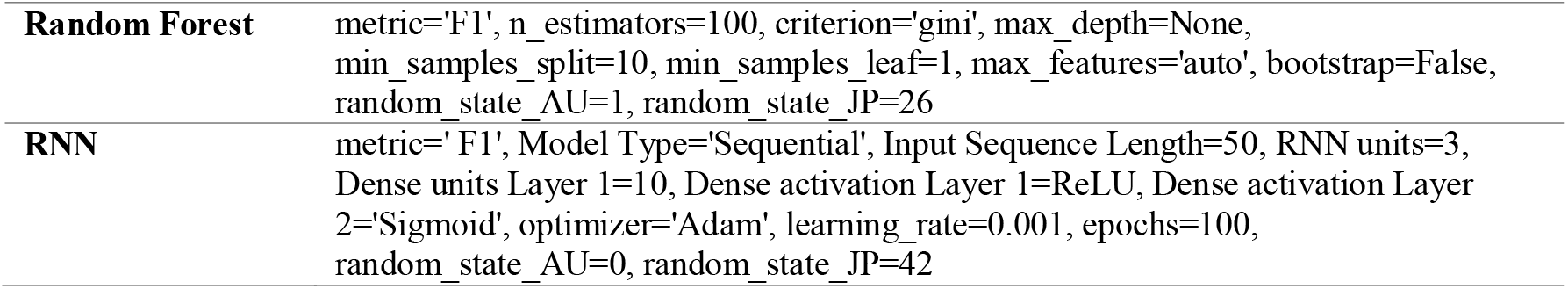
Hyperparameters of the algorithms

To ensure reproducibility and maintain data integrity across different cross-validation folds, an initial seed or state was set during data splitting. This guarantees, barring any noise, that simulations remain entirely deterministic based on the random seed.

The best overall performance, as measured by the ROC-AUC score, was achieved by the Random Forest algorithm (0.98, see Table 3). Nonetheless, this score was similar to the Decision Tree model, which exhibited exceptional performance on the Australian dataset, using stratified k-fold (k=5) cross-validation.

**Table 3.**
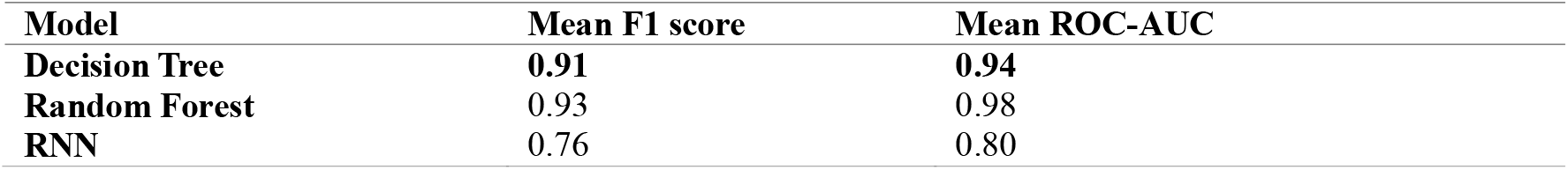
Performance metrics with the Australian Dataset

he model, pre-trained on the under-sampled Australian dataset with optimal hyperparameters, then underwent transfer learning for evaluation on the Japanese dataset. This involved fine-tuning or calibrating the model on a small 15% subset of the Japanese dataset, maintaining the same feature set before assessing its performance on the remainder of the data. The subset’s label distribution mirrored the original dataset, ensured by stratified k-fold sampling. Similarly to the internal validation using the Australian dataset, setting the initial seed or state during data splitting plays a crucial role in influencing the outcomes. **Table 4** contrasts the results of the transfer learning and evaluation process on the Japanese dataset in comparison with the Australian dataset.

**Table 4.** Performance comparison of all algorithms on the Australian and Japanese datasets.

|  | **Stratified K Fold (K=5)**<br>**Validation on AU Dataset**<br>**(Mean and Standard**<br>**Deviation)** | **Evaluation on JP**<br>**Dataset**<br>**(no fine tuning)** | **Evaluation on JP**<br>**Dataset**<br>**(15% fine tuning)** |
| --- | --- | --- | --- |
| **Decision Tree** |  |  |  |
| Accuracy | 0.907±0.035 | 0.752 | 0.860 |
| Precision | 0.918±0.047 | 0.597 | 0.691 |
| Recall | 0.895±0.030 | 0.265 | 0.876 |
| f1_score | 0.906±0.034 | 0.367 | 0.773 |
| Specificity | 0.918±0.050 | 0.933 | 0.854 |
| ROC-AUC | 0.941±0.031 | 0.750 | 0.882 |
| **Random Forest** |  |  |  |
| Accuracy | 0.926±0.009 | 0.738 | 0.878 |
| Precision | 0.928±0.008 | 0.571 | 0.750 |
| Recall | 0.923±0.021 | 0.148 | 0.826 |
| f1_score | 0.926±0.010 | 0.235 | 0.786 |
| Specificity | 0.929±0.009 | 0.958 | 0.897 |
| ROC-AUC | 0.976±0.004 | 0.794 | 0.928 |
| **RNN** |  |  |  |
| Accuracy | 0.753 ± 0.070 | 0.472 | 0.638 |
| Precision | 0.738 ± 0.074 | 0.331 | 0.422 |
| Recall | 0.801 ± 0.107 | 0.926 | 0.906 |
| f1_score | 0.764 ± 0.068 | 0.488 | 0.576 |
| Specificity | 0.705 ± 0.120 | 0.303 | 0.538 |
| ROC-AUC | 0.799 ± 0.065 | 0.615 | 0.794 |

The confusion matrices and ROC-AUC graphs are illustrated in **Figures 3-4**.

**Figure 3.**
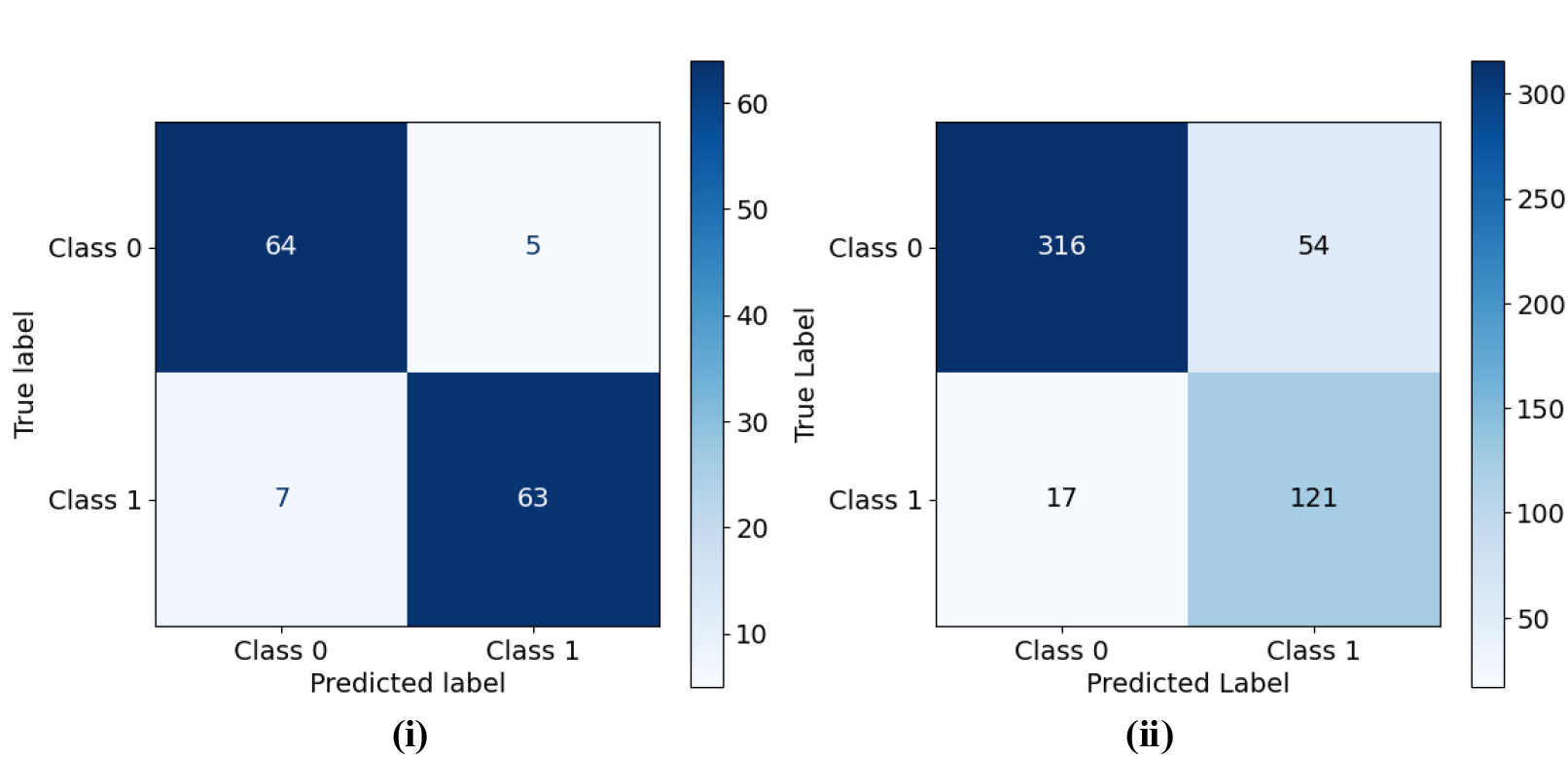
i) Confusion Matrix for the Decision Tree: Averaged Results across Stratified K Fold (K = 5) Validation on Australian Dataset; ii) Confusion Matrix for the Decision Tree: Evaluation on Japanese Dataset (15% fine tuning).

### Explainability

To further assess and corroborate with the findings from the statistical analyses, the Decision Tree model was additionally trained independently on the JP dataset, along with the AU dataset, utilising stratified K-Fold validation to discern predictive differences (see Table 5). Feature importance rankings for the AU dataset now show a significant prioritisation of eGFR (0.735), with eGFR_slope (0.172) and eGFR_mean (0.056) also contributing but to a lesser extent, followed by AGE (0.035) and SEX (0.002). For the JP dataset, the model similarly identified eGFR (0.450) as the most influential predictive feature, with eGFR_slope (0.297) and eGFR_mean (0.157) also important, whereas AGE (0.096), and SEX had no measurable impact. When the Decision Tree model was trained on the AU data and evaluated on the JP data with transfer learning, the feature importance reflected a similar hierarchy, with eGFR (0.455) remaining the most significant, followed closely by eGFR_slope (0.336) and then by eGFR_mean (0.131), AGE (0.078), and again SEX showing no discernible influence.

**Table 5.** Feature Ranking from Decision Tree

| Feature | AU Importance | JP Importance | Trained AU Evaluated JP |
| --- | --- | --- | --- |
| eGFR | 0.735 | 0.450 | 0.455 |
| eGFR_slope | 0.171 | 0.297 | 0.336 |
| eGFR_mean | 0.055 | 0.157 | 0.131 |
| AGE | 0.034 | 0.095 | 0.078 |
| SEX | 0.002 | 0.000 | 0.000 |

The prominence of the most recent eGFR in the AU cohort’s feature ranking underscores its critical role in outcome prediction, emphasising its significance as a key indicator of kidney function and its broad range and variability. Conversely, the JP dataset’s model indicates that the rate of change in eGFR, as captured by eGFR_slope, is comparatively more critical for predictions than for the AU cohort, potentially due to the dataset’s negative mean eGFR slope, which underscores the declining kidney function’s role in prognosis. Despite eGFR_mean’s reduced importance in the AU dataset compared to the JP dataset, it remained a relatively significant predictor, indicating nuanced differences in how eGFR trends impact prognosis across populations. Age held a more predictive power in the JP dataset than in the AU dataset, potentially reflecting the cohorts’ demographic variations in age. The negligible role of gender in both models, with no discernible influence in the JP dataset and only a marginal presence in the AU dataset, suggests that gender may have a limited direct influence on the prognostication of kidney function outcomes. When trained on the AU dataset and evaluated on the JP data, the model similarly emphasises eGFR and eGFR_slope, further highlighting the consistency of these features’ predictive importance across different population datasets. These variances in feature importance not only reveal intrinsic distinctions between the AU and JP datasets but also underscore the necessity for dataset-specific model tuning to enhance predictive performance in heterogeneous populations.

Figure 5. provides a detailed visualisation of the decision tree model trained on 85% of the Japanese dataset post the fine-tuning process using 15% of the data. This classification tree represents how the model discriminates between two outcomes based on a series of feature-based decisions.

**Figure 4.**
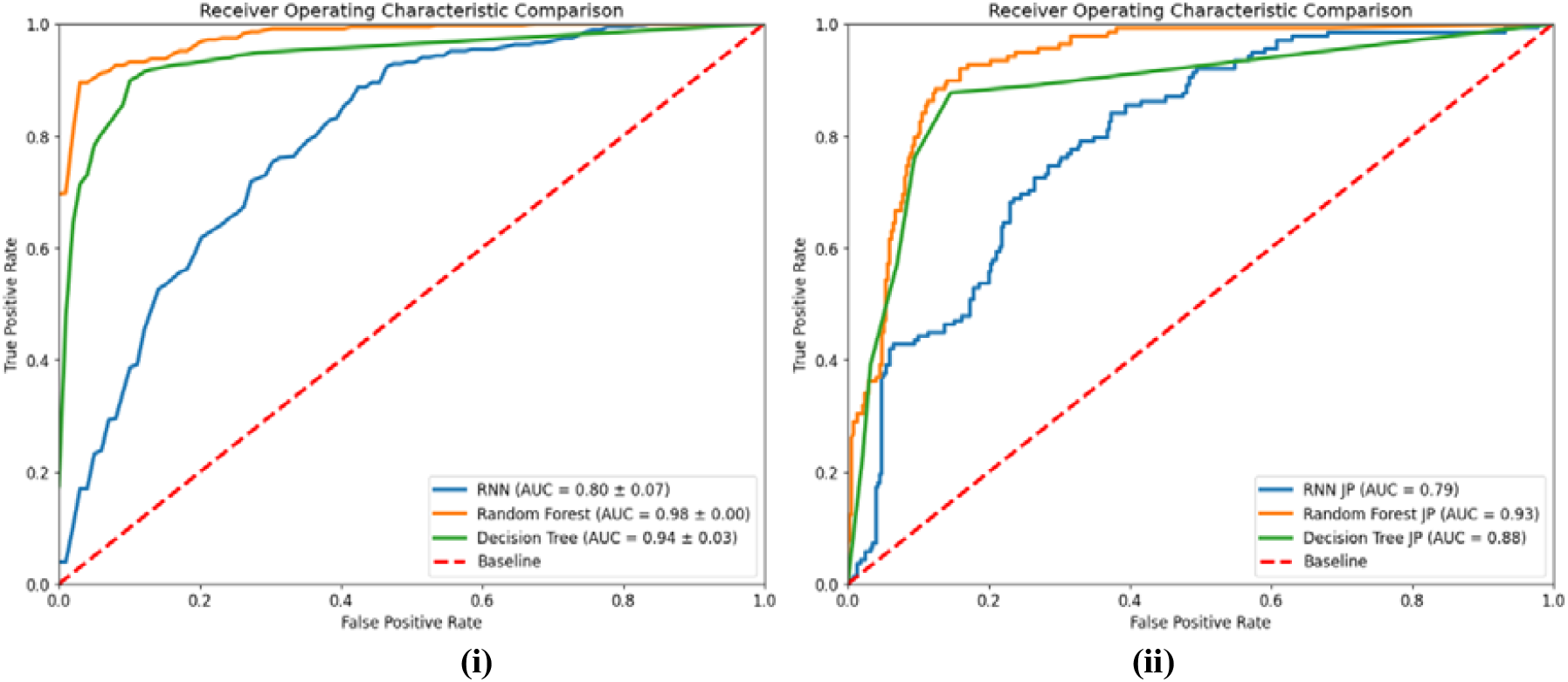
i) ROC-AUC Graph for All Comparative Models: Stratified K Fold (K = 5) Validation on Australian Dataset; ii) ROC-AUC Graph for All Comparative Models: Evaluation on Japanese dataset (15% fine tuning).

**Figure 5.**
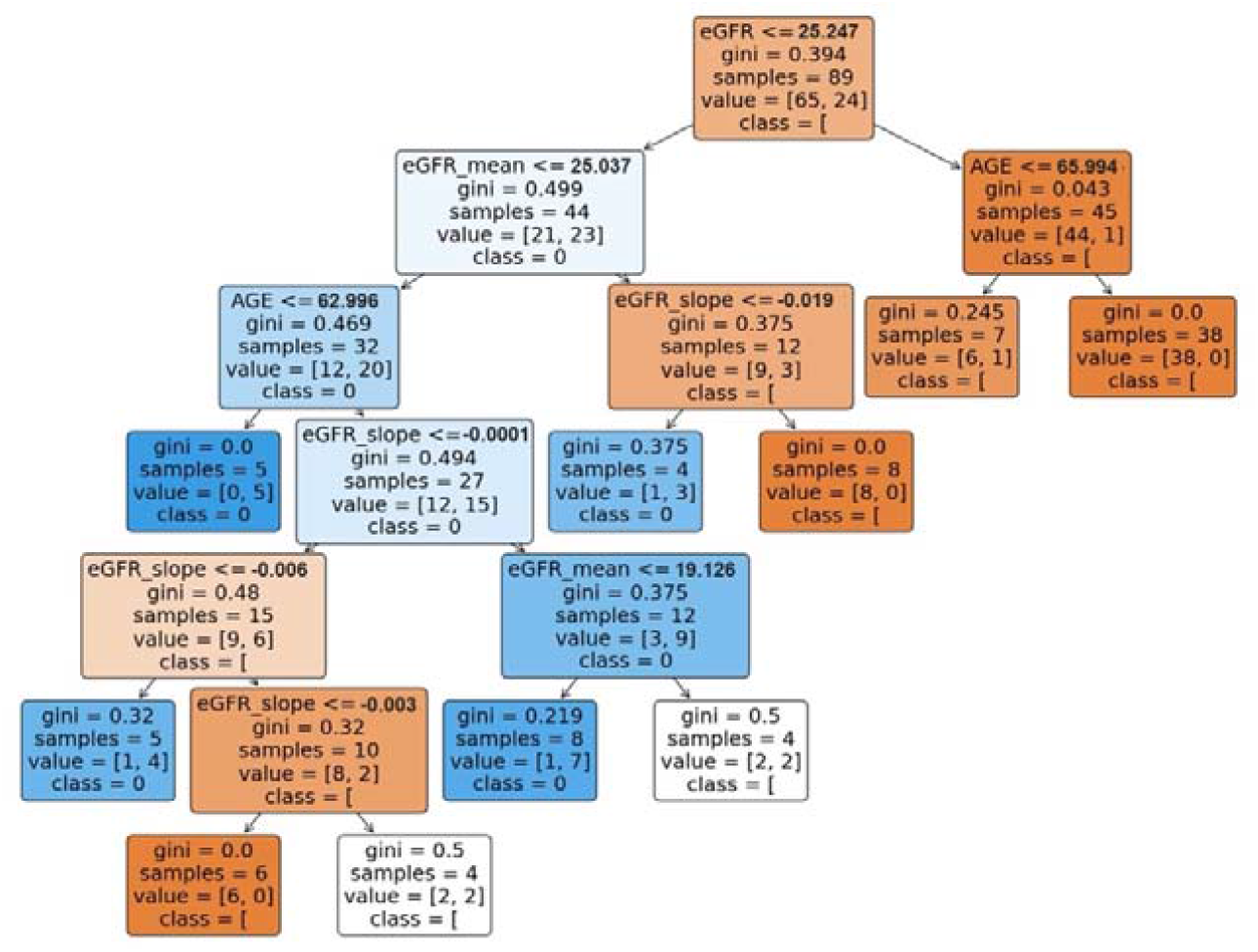
Decision Tree Rules (Cross-validation on Japanese dataset).

In the depicted decision tree, the primary attribute for initiating the division is eGFR, indicating its pivotal role in the model’s prognostications. Subsequent to the root node, the tree utilises additional divisions based on eGFR_slope and AGE, further emphasising their contribution to the classification process. Importantly, SEX does not manifest as a divisional variable at any of the principal nodes within this tree, underscoring its relatively minor significance in comparison to eGFR-related attributes and AGE. The tree structure delineates the decision-making pathway, revealing how the values of each attribute guide the prognostic outcomes. For example, the tree suggests that higher values of eGFR_mean lead to distinct classification outcomes, which may correspond to differing risks associated with renal function. This decision tree provides a transparent, interpretable model that elucidates the key factors driving the classification of renal function outcomes in the Japanese dataset. It delivers a direct and comprehensible perspective of the attribute relationships and their significance, rendering the tree’s logic and the impact of each variable on the outcomes readily apparent. The visual representation allows for an intuitive understanding of the model’s classification logic and the significance of each variable within the dataset.

### Decision Tree SHAP Interpretation

SHAP (Shapley Additive exPlanations) values measure the impact of each feature on the model’s prediction. For decision trees, SHAP can provide an exact solution for SHAP values, representing the average contribution of a feature value to every possible prediction. Figure 8 displays a SHAP summary plot. SHAP values explain the output of machine learning models by assigning each feature an importance value for a particular prediction. The SHAP summary plot revealed the relative importance of features in influencing a machine learning model’s prediction. The feature eGFR stands out as the most influential. The features eGFR_mean, eGFR_slope and AGE also play notable roles, whereas the SEX feature appears to have a more subdued impact on the model’s predictions overall. The visualisation in **Figure 6** underscores the importance of understanding individual feature contributions to enhance the model’s interpretability and trustworthiness. Then, across each feature, each dot represents the impact of a feature value on the model’s prediction for a single patient, with the position on the x-axis indicating the magnitude and direction of the impact. In the plot for the positive class, it is observed that higher eGFR values tend to decrease the risk of kidney failure (blue dots on the left side), while lower values increase the risk (pink dots on the right side). This aligns with medical understanding, as eGFR is a critical indicator of kidney function.

**Figure 6.**
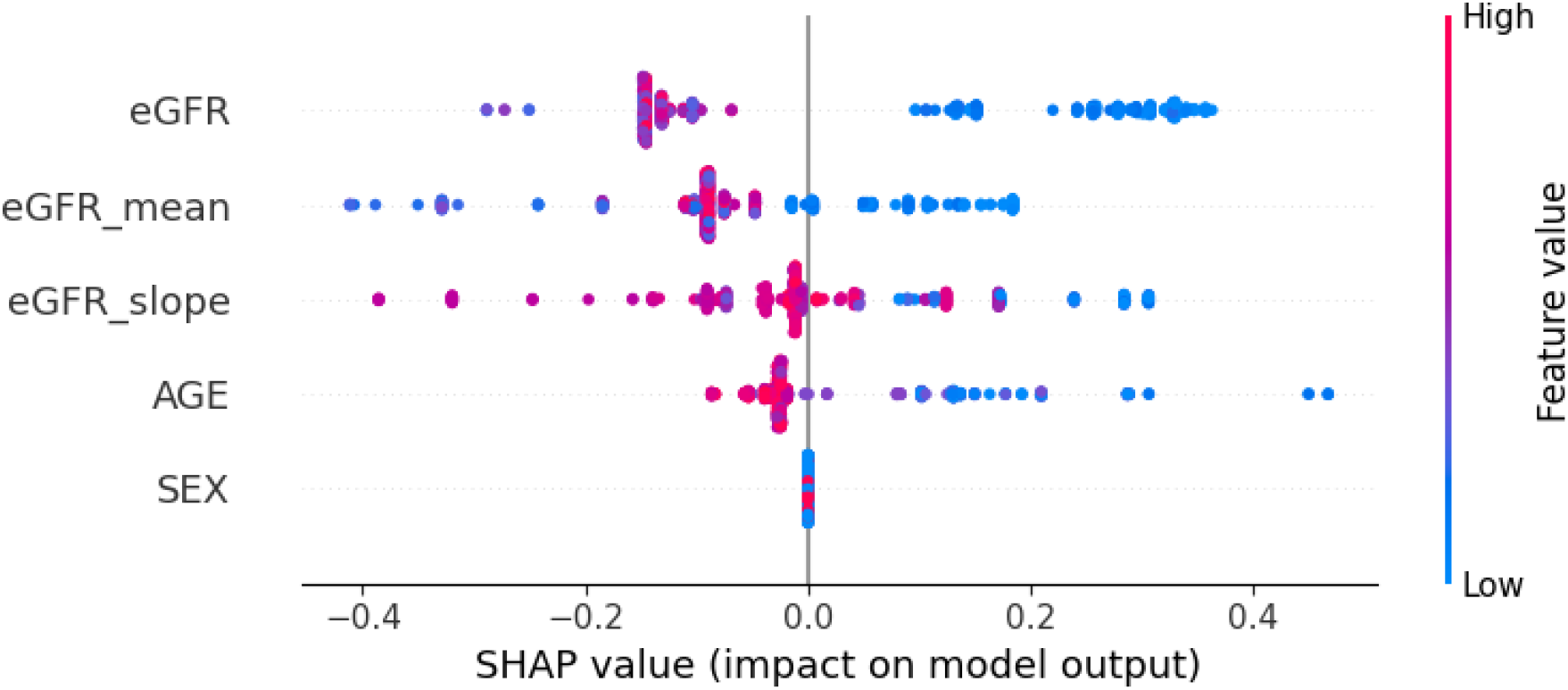
SHAP summary plot illustrating the global influence of input variables on the predictive performance of the Decision Tree model (cross-validation on Japanese dataset), using SHAP values.

### Decision Tree LIME Interpretation

Additionally, LIME analysis was applied to analyse and interpret the prediction for a particular instance. The LIME results offer insights into the local behaviour of a machine-learning model for a specific prediction. This analysis was illustrated by examining an individual patient, as depicted in **Figure 7**. The interpretive figure showcases how specific features sway the classification towards or against Kidney Failure, with orange indicating a positive influence and blue a negative one. The figure’s left side presents the prediction probabilities, and the central portion ranks the most influential features by bar length. For this instance, the model predicts “Kidney Failure” with absolute certainty. In the figure’s explanation, the eGFR_mean (15.36 mL/min/1.73m^2^) feature heavily favours a Kidney Failure outcome, followed by eGFR (15.33 mL/min/1.73m^2^) and AGE (27 years). The eGFR_slope (-0.0013) has a very minor influence, and SEX (male) has no impact on this prediction. This aligns with the clinical understanding that lower eGFR values are indicative of poorer kidney function, and the model has learned to heavily weigh these features when predicting Kidney Failure. The actual label for this case is also “Kidney Failure,” confirming the model’s prediction. In summary, the model’s prediction of “Kidney Failure” is strongly guided by the eGFR_mean, eGFR and AGE features. LIME clarifies these individual predictions, contrasting them with the broader, often opaque model behaviours. LIME’s strength lies in explaining individual predictions, rather than global model behaviour, by creating a local surrogate model that approximates the behaviour of the complex model around the vicinity of a particular instance. The surrogate model, often a linear model, is interpretable because it weights the input features in a way that can be easily understood.

**Figure 7.**
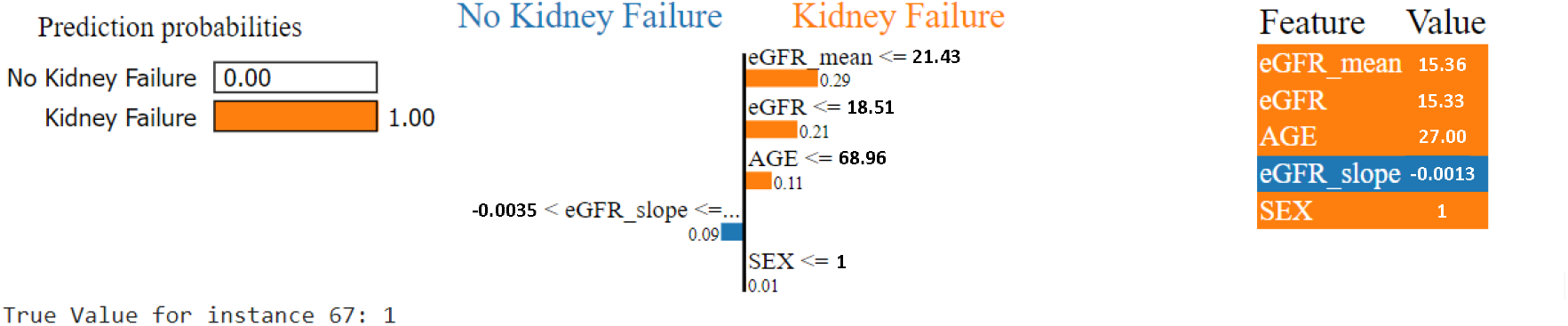
LIME plot illustrating the local influence of input variables on the predictive performance of the Decision Tree model (cross-validation on Japanese dataset) for a particular patient instance.

LIME can help understand the rationale behind specific decisions made by complex models.

This contrasts with more complex models, like deep neural networks or ensemble methods, which might be considered “black boxes” due to their intricate internal workings that are not readily interpretable.

### Counterfactual Analysis

Counterfactual analysis is an interpretability technique for understanding model predictions by exploring “what if” scenarios and querying changes required to impact the predicted outcomes. It involves generating counterfactual examples: slight alterations to the input features of a given instance that would lead to a different prediction outcome. This analysis is valuable for interpretability and explainability in machine learning, as it provides actionable insights into how feature values influence predictions, enhancing transparency and trust in complex models. DiCE (Diverse Counterfactual Explanations) is an interpretability framework designed to help understand ML model predictions by generating counterfactual explanations(19). These hypothetical scenarios show minimal changes needed in the input features of a data instance to achieve a different prediction outcome. Generating counterfactuals with DiCE involves finding instances in the feature space close to the original instance, which results in a different prediction. In the case of patient UID 67, which was predicted to have Kidney Failure, DiCE was used to find what changes could be made to this instance’s features to change the model’s prediction to Non-Kidney Failure. A counterfactual example that resulted in this was changing the eGFR_mean from 15.396 mL/min/1.73m^2^ to 25.483 mL/min/1.73m^2^, implying that increasing the average eGFR value slightly could also contribute to a Non-Kidney Failure prediction. Other features remain unchanged in this counterfactual example, highlighting that altering the eGFR alone, in this case, could lead to the desired prediction change. The counterfactuals suggest that small adjustments to eGFR, eGFR_mean, and eGFR_slope could lead to a prediction of non-Kidney failure for this patient. These changes are hypothetical and need to be interpreted within the clinical context. For example, an increase in eGFR values generally indicates better kidney function, which aligns with the counterfactual prediction of non-Kidney failure. It is important to note that these counterfactuals do not imply causation but provide potential scenarios that could lead to a different prediction, which can be useful for understanding model behaviour and sensitivities. The effectiveness and plausibility of these counterfactuals depend on the accuracy and reliability of the model and the representativeness of the data it was trained on. In healthcare applications, any insights or actions derived from such analyses should be evaluated and validated by medical professionals. This analysis could extend to more personalised and alterable variables such as BMI, smoking and other lifestyle factors.

## Discussion

Early identification of individuals at risk of progressing to kidney failure is crucial, as it allows for targeted interventions that can significantly alter the course of the disease (5). Although CKD progresses through each stage sequentially, it is often clinically silent, meaning that most affected individuals are unaware of their condition (5, 7). As a result, many are diagnosed at mid to late stages, necessitating urgent treatment. The early methods for predicting CKD progression involve using statistical methods - the most sophisticated currently being the KFRE (11). Although this equation provides great insight into the kidney failure detection, it is a static equation which focus only on the current values and does not take the time-series changes into consideration for kidney failure prediction. The advent of AI has brought about a revolutionary shift in data handling and processing (20). Although the fundamental calculations remain consistent, the scale and predictive capabilities have entered a completely new realm. Traditional methods can be both expensive and time-consuming, leaving greater margin for human error; thus, machine learning offers promising solutions for these reasons. Machine learning has made it feasible to extract meaningful insights from what would otherwise be inscrutable data, capturing underlying patterns and fluctuations.

Therefore, machine learning is better suited than traditional statistical models when dealing with non-linear data.

The current literature suggests a keen interest in the development of effective clinical utility prognostic model with the ability to predict kidney failure (10, 16). To achieve this, larger and more time-series data is being used for model training. Using machine learning for time-based variables rather than traditional regression models allows it to capture complex non-linear relationships. This cost-efficient, generalisable tool helps predict progressive CKD earlier across all disease stages compared to previous kidney failure prediction models.

The literature has also emphasised that the explainability of the CKD predictive model is necessary in a clinical environment (21). Training models from time-series data enable more complex, non-linear relationships between variables without resorting to ‘black-box’ methods such as deep neural networks where explainability is often a major difficulty. A deeper understanding of CKD over time can allow professionals in clinical settings to study disease evolution, paving the path to more personalised and specific medicine approaches. Various studies have taken the initiative and have expressed the need to build models based on time-varying data set (10, 16). It is also observed that studies have started to adopt more interpretable methods (14), recognising that the clinical practice requires a certain degree of interpretability for practical implementation. Our study has employed techniques such as LIME and SHAP, which provide a detailed analysis of the model’s decision-making process and rationale, offering insights at the local level (for an individual patient) and at the global level (across the entire dataset). This analysis helps in understanding the predictive behaviour of the model, including the identification of key features and impact on predictions, thereby offering a comprehensive view of how the model arrives at its conclusions in a wide range of scenarios. Utilizing counterfactual analysis, we further explore ‘what-if’ scenarios, examining the impact of the smallest changes in input variables on the model’s predicted risk of kidney failure.

Our retrospective cohort study demonstrates the potential utility of explainable machine learning approaches in enhancing the prediction of chronic kidney disease (CKD) progression. The results highlight exceptional predictive accuracy, including AUC-ROC scores of 0.94 and 0.98 for the decision tree and random forest models, respectively, in the Australian dataset. Following a transfer learning approach to enable validation in the smaller Japanese dataset of just 597 patients, AUC-ROC scores of 0.88 and 0.93 were achieved for the decision tree and random forest models, respectively. The high level of performance exhibited by these interpretable models underscores their ability to predict CKD outcomes even on limited sample sizes, while also providing clinical interpretability. The robust AUC-ROC values in our study are consistent with previous studies demonstrating the capabilities of machine learning for prognostic modelling in CKD. Our finding of comparable high accuracy for the random forest model aligns with these results and establishes machine learning techniques as valuable tools for risk-stratifying CKD patients.

Furthermore, the use of transfer learning, though novel in the CKD domain, enabled the validation of our models on the smaller Japanese dataset while retaining accuracy. Transfer learning involves transferring knowledge gained from training a model on a source dataset to improve performance on a different target dataset. This technique has shown tremendous promise for medical imaging, genomic analyses, and other biomedical applications with limited data (22). By leveraging the Australian discovery cohort to pre-train, the decision tree and random forest models before fine-tuning and evaluation on the Japanese dataset, we mitigated overfitting issues and demonstrated wider applicability across ethnically diverse populations.

The feature importance analysis also provides clinical interpretability by revealing the relative influence of patient age, most recent eGFR, eGFR slope, and mean eGFR on outcome predictions by the models. Understanding the impact of these variables can inform targeted early interventions to mitigate risk. For example, increased monitoring and renoprotective therapies could be implemented for patients exhibiting a sharply declining eGFR over time. Previous studies have similarly called for explainable and interpretable machine learning models in CKD to enable actionable insights into patient trajectories (5,6).

Several limitations temper the results and warrant discussion. The modest sample sizes, though partly addressed via transfer learning, necessitate validation in considerably larger independent cohorts. A large and geographically diverse outpatient cohort would have strengthened its overall generalizability and usability. Additionally, the consistency of the models over longer follow-up was not evaluated. Furthermore, seamless integration with clinical workflow, including EHR systems, would be essential to enable pragmatic implementation. Finally, additional renal and cardiometabolic parameters could be incorporated to enhance risk stratification capabilities.

Our findings provide preliminary evidence supporting the integration of novel prognostic variables and interpretable machine learning techniques to predict early CKD progression. Some machine-learning models rely primarily on novel kidney disease biomarkers (23), while our models use readily obtainable laboratory information.The high predictive accuracy and transportability exhibited by the decision tree and random forest models position them as potentially valuable augmentations to existing tools like KFRE. Following external validation, such models could assist clinicians by identifying patients at high-risk of progression and who would likely benefit from close monitoring and treatment escalation. Future studies should implement these methods prospectively over extended periods in diverse CKD populations.

## Conclusion

In conclusion, this study has successfully developed explainable machine learning models utilising routinely collected pathology data to accurately predict the trajectory of CKD. By leveraging key variables such as age, gender, and various aspects of estimated eGFR, the models demonstrated exceptional predictive accuracy, particularly in identifying the risk of progression to kidney failure. Internal validation on an Australian cohort showcased high performance, with area under the receiver operating characteristic curve (ROC-AUC) values of 0.94 and 0.98 for decision tree and random forest algorithms, respectively. Furthermore, external validation on a Japanese dataset, followed by transfer learning techniques to enhance generalisation, yielded respectable accuracy scores, indicating the potential applicability of the models across diverse populations. Notably, the identified variables align with the underlying pathophysiology of CKD, emphasising the clinical relevance of the developed models. Additionally, interpretable ML techniques such as SHAP and LIME for further analysis provided insights into model behaviour at both local and global levels. These methodologies contributed coherent and rational explanations, thereby enhancing the model’s overall transparency. While further validation in larger multi-ethnic cohorts is warranted to strengthen generalizability, the study’s findings underscore the promise of accurate predictive modelling in identifying high-risk CKD patients for personalised disease management interventions.

## Code Availability

The code for the machine learning models is available at https://github.com/roysup/Chronic-Kidney-Disease

## Data Availability

All data produced in the present study are available upon reasonable request to the authors

https://github.com/roysup/Chronic-Kidney-Disease

## Appendices

### I All hyperparameters for classifiers

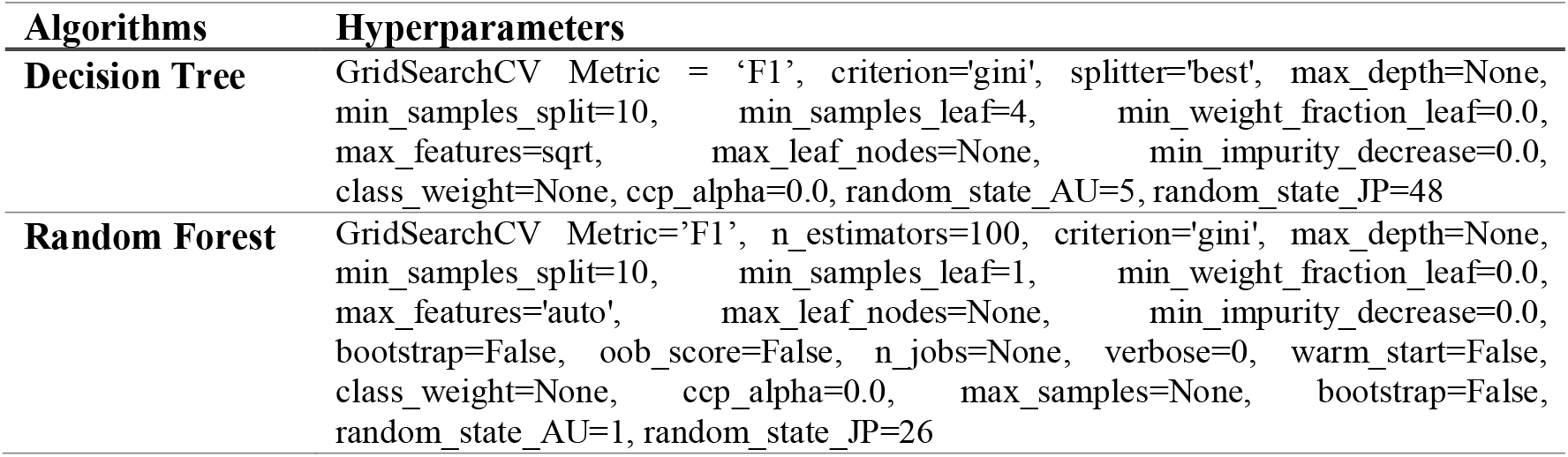
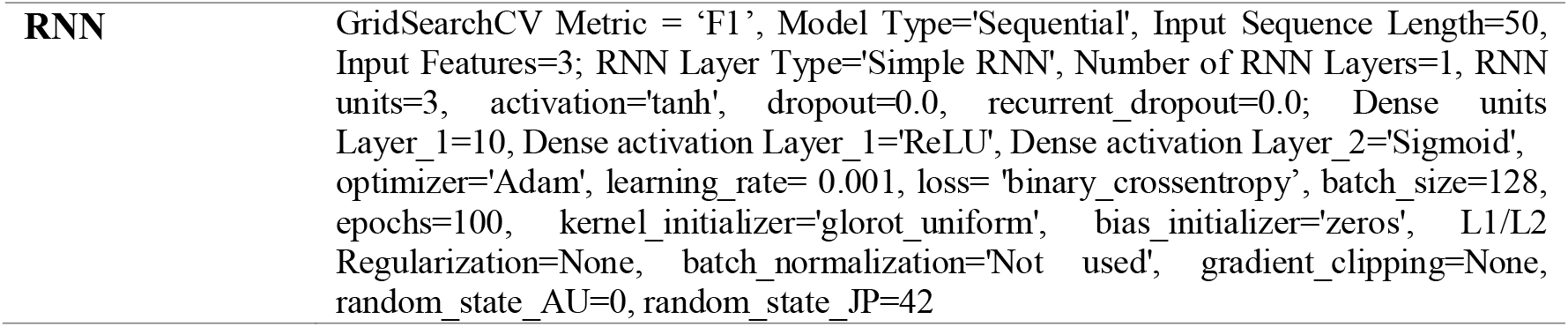

### II Decision Tree Rules for Predicting Kidney Failure: Pre-trained on Australian Dataset and Cross-validated on Japanese Population Data

~~~
if eGFR <= 25.24:
  if eGFR_mean <= 25.04:
    if AGE <= 63.00:
      return class = 1, distribution = [[0.0, 5.0]]
    else: # if AGE > 63.00
      if eGFR_slope <= -0.00:
        if eGFR_slope <= -0.01:
          return class = 1, distribution = [[1.0, 4.0]]
        else: # if eGFR_slope > -0.01
          if eGFR_slope <= -0.00:
            return class = 0, distribution = [[6.0, 0.0]]
          else: # if eGFR_slope > -0.00
            return class = 0, distribution = [[2.0, 2.0]]
      else: # if eGFR_slope > -0.00
        if eGFR_mean <= 19.13:
          return class = 1, distribution = [[1.0, 7.0]]
        else: # if eGFR_mean > 19.13
          return class = 0, distribution = [[2.0, 2.0]]
   else: # if eGFR_mean > 25.04
     if eGFR_slope <= -0.02:
       return class = 1, distribution = [[1.0, 3.0]]
     else: # if eGFR_slope > -0.02
       return class = 0, distribution = [[8.0, 0.0]]
 else: # if eGFR > 25.24
   if AGE <= 66.00:
     return class = 0, distribution = [[6.0, 1.0]]
   else: # if AGE > 66.00
     return class = 0, distribution = [[38.0, 0.0]]
~~~

### III Random Forest Decision Rules for Predicting Kidney Failure: Analysis of 10 Estimators Pre-trained on Australian Dataset and Cross-validated on Japanese Population Data

`Tree 1 Rules:`

~~~
if eGFR <= 22.49:
  if SEX <= 1.50:
   if  eGFR_mean <= 22.88:
     if eGFR_slope <= 0.00:
       return class = 1, distribution = [[3.0, 13.0]]
     else:  # if eGFR_slope > 0.00
       return class = 0, distribution = [[1.0, 0.0]]
     else:  # if eGFR_mean > 22.88
       return class = 0, distribution = [[6.0, 0.0]]
   else:  # if SEX > 1.50
     return class = 1, distribution = [[0.0, 12.0]]
 else:  # if eGFR > 22.49
   if eGFR_mean <= 25.10:
     return class = 1, distribution = [[0.0, 1.0]]
   else:  # if eGFR_mean > 25.10
     if AGE <= 53.50:
       return class = 0, distribution = [[1.0, 1.0]]
     else:  # if AGE > 53.50
       return class = 0, distribution = [[51.0, 0.0]]
~~~

`Tree 2 Rules:`

~~~
if eGFR_slope <= -0.02:
  return class = 1, distribution = [[0.0, 13.0]]
else:  # if eGFR_slope > -0.02
  if AGE <= 45.50:
    return class = 1, distribution = [[0.0, 3.0]]
  else:  # if AGE > 45.50
    if eGFR_slope <= -0.01:
      return class = 0, distribution = [[8.0, 4.0]]
    else:  # if eGFR_slope > -0.01
      if eGFR_slope <= -0.00:
        return class = 0, distribution = [[27.0, 0.0]]
      else:  # if eGFR_slope > -0.00
        if eGFR_mean <= 24.41:
          return class = 1, distribution = [[1.0, 4.0]]
        else:  # if eGFR_mean > 24.41
          return class = 0, distribution = [[29.0, 0.0]]
~~~

`Tree 3 Rules:`

~~~
if eGFR_slope <= -0.02:
  return class = 1, distribution = [[0.0, 10.0]]
else:  # if eGFR_slope > -0.02
  if eGFR_mean <= 22.39:
    if eGFR_slope <= -0.00:
      return class = 0, distribution = [[7.0, 2.0]]
    else:  # if eGFR_slope > -0.00
      return class = 1, distribution = [[1.0, 4.0]]
  else:  # if eGFR_mean > 22.39
    return class = 0, distribution = [[65.0, 0.0]]
~~~

`Tree 4 Rules:`

~~~
if eGFR_mean <= 25.10:
  if eGFR_slope <= -0.00:
    if AGE <= 79.00:
      if eGFR_mean <= 20.80:
        return class = 1, distribution = [[0.0, 7.0]]
      else:  # if eGFR_mean > 20.80
        return class = 0, distribution = [[6.0, 3.0]]
    else:  # if AGE > 79.00
      return class = 0, distribution = [[7.0, 0.0]]
  else:  # if eGFR_slope > -0.00
    return class = 1, distribution = [[0.0, 7.0]]
  else:  # if eGFR_mean > 25.10
     if AGE <= 53.50:
       return class = 0, distribution = [[3.0, 2.0]]
     else:  # if AGE > 53.50
       return class = 0, distribution = [[54.0, 0.0]]
~~~

`Tree 5 Rules:`

~~~
if eGFR <= 23.23:
  if AGE <= 75.50:
    if AGE <= 60.00:
      return class = 1, distribution = [[0.0, 11.0]]
    else: # if AGE > 60.00
      if eGFR_mean <= 22.75:
        if AGE <= 65.50:
          return class = 0, distribution = [[1.0, 0.0]]
        else: # if AGE > 65.50
          return class = 1, distribution = [[0.0, 14.0]]
      else: # if eGFR_mean > 22.75
        return class = 1, distribution = [[2.0, 3.0]]
   else: # if AGE > 75.50
     return class = 0, distribution = [[9.0, 5.0]]
  else: # if eGFR > 23.23
    if eGFR_slope <= -0.04:
      return class = 1, distribution = [[0.0, 2.0]]
    else: # if eGFR_slope > -0.04
      return class = 0, distribution = [[42.0, 0.0]]
~~~

`Tree 6 Rules:`

~~~
if eGFR_mean <= 25.86:
  if eGFR_slope <= 0.00:
    if AGE <= 79.50:
      if SEX <= 1.50:
        if eGFR_slope <= -0.00:
          return class = 1, distribution = [[5.0, 6.0]]
        else: # if eGFR_slope > -0.00
          return class = 1, distribution = [[0.0, 5.0]]
      else: # if SEX > 1.50
        return class = 1, distribution = [[0.0, 8.0]]
    else: # if AGE > 79.50
      return class = 0, distribution = [[4.0, 1.0]]
  else: # if eGFR_slope > 0.00
    return class = 0, distribution = [[4.0, 0.0]]
else: # if eGFR_mean > 25.86
  if AGE <= 63.50:
    return class = 0, distribution = [[7.0, 5.0]]
  else: # if AGE > 63.50
    return class = 0, distribution = [[44.0, 0.0]]
~~~

`Tree 7 Rules:`

~~~
if eGFR <= 20.25:
  if eGFR_mean <= 19.99:
    return class = 1, distribution = [[0.0, 11.0]]
  else: # if eGFR_mean > 19.99
    if eGFR_slope <= -0.02:
      return class = 1, distribution = [[0.0, 4.0]]
      else: # if eGFR_slope > -0.02
        return class = 0, distribution = [[6.0, 1.0]]
   else: # if eGFR > 20.25
     if AGE <= 53.50:
       return class = 0, distribution = [[2.0, 1.0]]
      else: # if AGE > 53.50
       if eGFR_mean <= 22.88:
         return class = 1, distribution = [[0.0, 1.0]]
       else: # if eGFR_mean > 22.88
         return class = 0, distribution = [[63.0, 0.0]]
~~~

`Tree 8 Rules:`

~~~
if eGFR_slope <= -0.01:
  return class = 1, distribution = [[1.0, 12.0]]
else:  # if eGFR_slope > -0.01
  if eGFR <= 19.54:
    if eGFR_slope <= -0.00:
      return class = 0, distribution = [[2.0, 0.0]]
    else:  # if eGFR_slope > -0.00
      if eGFR <= 16.39:
        return class = 1, distribution = [[0.0, 7.0]]
     else:   # if eGFR > 16.39
       return class = 1, distribution = [[2.0, 5.0]]
 else:  # if eGFR > 19.54
   if AGE <= 76.50:
     return class = 0, distribution = [[36.0, 0.0]]
   else:  # if AGE > 76.50
     if eGFR <= 24.82:
       return class = 0, distribution = [[2.0, 1.0]]
     else:  # if eGFR > 24.82
       return class = 0, distribution = [[21.0, 0.0]]
~~~

`Tree 9 Rules:`

~~~
if eGFR_mean <= 22.39:
  if eGFR_slope <= -0.01:
    return class = 1, distribution = [[0.0, 6.0]]
  else:  # if eGFR_slope > -0.01
    if eGFR_mean <= 16.88:
      return class = 1, distribution = [[0.0, 3.0]]
    else:  # if eGFR_mean > 16.88
      if AGE <= 79.50:
        return class = 1, distribution = [[2.0, 6.0]]
      else:  # if AGE > 79.50
        return class = 0, distribution = [[13.0, 1.0]]
else: # if eGFR_mean > 22.39
  if eGFR_slope <= -0.03:
    return class = 1, distribution = [[0.0, 5.0]]
  else:  # if eGFR_slope > -0.03
    if eGFR_mean <= 25.10:
      return class = 0, distribution = [[5.0, 1.0]]
    else:  # if eGFR_mean > 25.10
      return class = 0, distribution = [[47.0, 0.0]]
~~~

`Tree 10 Rules:`

~~~
if AGE <= 47.00:
  return class = 1, distribution = [[0.0, 2.0]]
else: # if AGE > 47.00
  if eGFR_mean <= 17.31:
    return class = 1, distribution = [[0.0, 12.0]]
  else: # if eGFR_mean > 17.31
    if eGFR_slope <= -0.02:
      return class = 1, distribution = [[0.0, 5.0]]
    else: # if eGFR_slope > -0.02
      if eGFR <= 19.68:
        return class = 0, distribution = [[12.0, 1.0]]
      else: # if eGFR > 19.68
        return class = 0, distribution = [[57.0, 0.0]]
~~~

The rules provided from the singular decision tree and each estimator in the Random Forest model give a nuanced view of how different features contribute to predicting kidney failure (class 1). Here are some insights and considerations:

- Low eGFR values: Across multiple trees, lower values of eGFR often lead to predicting kidney failure (class 1), which aligns with medical knowledge. eGFR (estimated Glomerular Filtration Rate) is a critical measure of kidney function, and lower values indicate poorer kidney function.
- eGFR_mean and eGFR_slope: These features, representing the mean eGFR value and the slope of eGFR over time, respectively, are crucial in predicting kidney failure. For example, a more negative eGFR_slope (indicating a faster decline in kidney function) often leads to predictions of kidney failure.
- Impact of AGE and SEX: The rules also consider AGE and SEX in making predictions. It’s noteworthy that in some trees, these features directly influence the prediction, highlighting their importance in the model’s decision-making process. For instance, in Tree 1, if eGFR is above a threshold and SEX is greater than 1.50 (1: Male; 2: Female), it predicts class 1, suggesting that gender might have a significant role in the context of other features.
- Distribution in leaf nodes: The class distributions in the leaf nodes (e.g., [3.0, 13.0]) give an idea of the model’s certainty. A more balanced distribution (e.g., [1.0, 1.0]) suggests less certainty, while a skewed distribution (e.g., [0.0, 12.0]) indicates higher confidence in the prediction.
- Variability across trees: The diversity in rules and thresholds across different trees exemplifies the strength of Random Forests in capturing complex patterns through ensemble learning. Each tree might be capturing different aspects or interactions of the features.
- Clinical relevance: The thresholds for features like eGFR, eGFR_mean, and eGFR_slope should be evaluated in the context of clinical relevance. Their values and the model’s reliance on them should make sense from a medical perspective for diagnosing kidney failure.

In summary, the rules from these trees seem to make sense for predicting kidney failure, given the known importance of eGFR as a kidney function marker. However, the effectiveness and reliability of these predictions should also be validated with medical experts to ensure they align with clinical knowledge and practice.

